# A qualitative and quantitative analysis of changes in prostate MRI T2WI signals with different abstinence durations

**DOI:** 10.1101/2024.05.03.24306819

**Authors:** Wenjun Ma, Baoming Ren, Yanjun Gao, Weixian Bai

**Affiliations:** Department of Urology, XI’an No.3 Hospital, the Affiliated Hospital of Northwest University, Xi’an, Shaanxi 710018, P.R. China; Department of Medical Imaging, XI’an No.3 Hospital, the Affiliated Hospital of Northwest University, Xi’an, Shaanxi 710018, P.R. China

**Author notes:** Corespongding author. Co-first author: Wenjun MA, Baoming REN.

## Abstract

**PURPOSE:** To investigate the effect of abstinence duration on image quality of prostate with high-field magnetic resonance imaging(MRI);

**MATERIALS AND METHODS:** This study included males who underwent prostate MRI in Xi’an No.3 Hospital from November 2021 to November 2022.And these males were divided into two groups according to whether they had ejaculation before MRI examination within 3 days. Two radiologists blinded evaluated the boundary sharpness of the peripheral zone (PZ) and central gland (CG) of prostate using a three-point Likert-scale. The region of interests (ROIs) of the PZ and CG at both sides on the sagittal T2W image were drawn and the signal intensity (SI) of ROIs was measured, then the SI ratio (SIR) was calculated. The Wilcoxon’s test and the independent-samples T test was utilized to compare the differences of the two groups;

**RESULTS:** A total of fifty young males were included in the study. There were twenty males abstinence more than 3-days, and thirty males were abstinence within 3-days. The qualitative analysis showed that the different structure of prostate in abstinence group was easier to identify than in ejaculation group(P≤0.05). The quantitative analysis showed that the SIR of PZ and CG in the ejaculation group were lower than in the abstinence group (all *P*≤0.05), and the peripheral zone was significantly lower;

**CONCLUSION:** In order to make the results more accurate, we suggested that the abstinence should more than 3 days prior to the examination.

## INTRODUCE

Prostate cancer (PCa ) is one of the most common cancer of male, at world-widely^[1-2]^. In China, it is the sixth-most common cancer of male and the prevalence is increasing in recent years ^[3-4]^. MRI played an important role in assessment of PCa, which is widely used as a tool of diagnosis, staging, judgment prognosis, and provide evidence for clinical decision making ^[5-7]^. The Prostate Imaging-Reporting and Data System version 2.1(PI-RADS v2.1) was published in 2019, it has been validated in clinical work and study ^[8]^. To make sure the quality of management, communication and multicenter of PCa, the PI-RADS v2.1 formulates specifications for the acquisition technology and interpretation of prostate MR images. Multi-parameter MRI (mp-MRI), include T2-weighted imaging (T2WI), diffusion-weighted imaging (DWI) and dynamic contrast-enhanced MRI (DCE-MRI), was recommended as the first-line MR sequences.

In some culture, sexual activities in the elderly people are considered to be uncommon or even strange. As previous studies have shown that, for the majority elder males, sexual activities palyed an important role. In a research about age-dependent frequency of sexual activities in Germany,in age groups between 60 and 79 years, the frequency of sexual activity over the months prior to the study was between 71% and 84%. In Japan, the frequency of same age group was 55%-88%. Therefore, the sexual activity of elder people cannot be ignored.

Several previous MRI studies reported that the T2 valued and apparent diffusion coefficient (ADC) value of prostate changed dynamically with ejaculation ^[9].^ PI-RADS v2.1 mentioned that “some recommend that patients refrain from ejaculation for three days prior to the MRI exam”, but it also high-lighted that “the benefit of abstinence has not been firmly established” ^[10].^ Thus the purpose of this study was to retrospective analyze high-filed T2WI of prostate with different abstinence time through qualitative and quantitative method, to investigate the influence of abstinence to the prostate T2WI, and to determine whether should be abstinence for 3 days.

## MATERIALS AND METHODS

### Study population

A total number of 128 healthy males had underwent prostate MRI examination from November 2021 to November 2022. But 78 patients were excluded: 34 were lack of detailed clinical records or MR imaging without T2W-images,31 were younger than 20 years or older than 35 years, and 13 patients for the reason of poor image quality. Finally, 50 patients were enrolled in our study. All of the patients had T2-weighted imaging (both axial- and sagittal-images) and reliable abstinence time. According to the abstinence time, the patients were divided into two groups, abstinence group abstained for more than 3 days (abstinence, n=20,60%), and ejaculation group had ejaculation within 3 days (ejaculation, n=30, 40%). Our retrospective study was approved by the hospital ethics committee of Xi’an NO.3 Hospital. And All patients were informed and signed informed consent forms.

### MRI acquisition

MRI of the prostate were performed on a 3-T MR scanner (Philips Ingenia, Philips Healthcare, Best, The Netherlands) with an 18-channel phased-array body coil and a 32-channel phased-array spine coil. Three-dimensional (3D) high-resolution T2-weighted images were acquired using a 3D turbo spin echo (TSE) with the following parameters: TR/TE= 3556/113 ms/ms, field of view (FOV) = 180*180mm^2^, Slice thickness 3 mm, Gap 0 mm.

### MRI image analysis

The images were saved and transferred in DICOM(Digital Imaging and Communications in Medicine ) format and patient information was hidden. All high-resolution T2-weighted images were transferred to post-processing workstation and display the axial and sagittal images of the prostate.

### Qualitative analysis

Two urogenital radiologists, respectively with 5-7 years of experience in prostate MRI, randomly reviewed the images of patients. The readers independently evaluated the boundary between the central gland and peripheral zone on axial and sagittal T2-weighted images based on a three-point Likert-scale (3, well-defined; 2, obscure; 1, ill-defined). All the process of evaluation was supervised by a senior radiologist. When the opinion was different, discussed with senior radiologist and determined the finally grade.

### Quantitative analysis

ROIs were drew and analyzed by a radiologist with 5 years of experience in prostate MRI through ITK-SNAP software((ITK-snap v.3.6.0, http://www.itksnap.org), and supervised by the senior radiologist. The T2-weighted signal intensity was measured by drawing four cycle-shaped ROIs within prostate at sagittal position, the diameter of ROI is 1 cm, respectively represent PZ and CG on both symmetrically layer. And also drawing a same size ROI in obturator internus. Then, the signal intensity ratio (SIR) of each ROIs was calculated with the signal intensity of obturator muscle as denominator.

### Statistical analysis

Statistical analysis was performed using R version 3.1 (The R Foundation for Statistical Computing). The Likert-scale were expressed as mean with minimum and maximum value. The Wilcoxon’s test was utilized to compare the differences of Likert-scale between the two groups. The SIR was presented as the mean(standard deviation,SD). We used the Shapiro-Wilk test to confirm normality of the data distribution. The independent-samples T test was used to analyze normally distributed data, and Wilcoxon’s test was applied to non-uniform distributed data. A *p*-value<0.05 was considered to be statistically significant.

## RESULTS

### Patient characteristic

There were 20 males (40%) in abstinence group, the mean age was 27.55 years old (range from 22.7-34.6), the mean abstinence time was 9.13 days (range from 5-15); And 30 males (60%) in ejaculation group, the mean age was 28.15 years old (range from 22.5-34.9), the mean abstinence time was 2.13 days (range from 1-3). There was significant difference in abstinence time between the two groups (*P*=0.000), but no difference in age (*P*=0.772) (Table 1) .

**Table 1.** Patient characteristic of two groups.

| **Group** | **n** | **Age** | **Time** |
| --- | --- | --- | --- |
| **Abstinence** | 20 | 27.55(4.27) | 9.13(3.52) |
| **Ejaculation** | 30 | 28.15(3.85) | 2.13(0.83) |
| **P-value** | — | 0.772 | 0.000 |

*Data are presented as mean (SD).*

### Qualitative analysis

The sharpness of boundary between the central gland and peripheral zone showed difference between the two groups (*P*=0.000). There are 50.00%(10/20) of T2-images in abstinence group were well-defined, only 10.00% (2/20) were ill-defined. On the contrary,most (70.00%, 21/30) T2-images in ejaculation group were hard to define the boundary between the two structures, there are only (3.33%, 1/30)were well-defined(Table 2).

**Table 2.** The sharpness of boundary between the central gland and peripheral zone in two groups.

| Group | Score |  |  |
| --- | --- | --- | --- |
|  | Ill-defined | Obscure | Well-defined |
| Abstinence | 2 | 8 | 10 |
| Ejaculation | 21 | 8 | 1 |
| $\chi^2$ | | 21.937 | |
| $P$-value | | 0.000 | |

### Quantitative analysis

In the same group, to compare the SIR of the central gland (CG), the peripheral zone (PZ) signal ratio (SIR), and the PZ-CG between left and right sides, and there are no difference between the left and right sides in the same group (Table 3, Fig1.). The SIR of CG, PZ and PZ-CG on the same side between the two groups were statistically different (all *p*<0.05) (Fig 2.).

**Table 3.** The SIR at different ROI of both side in two groups.

|  | Left |  |  | Right |  |  |
| --- | --- | --- | --- | --- | --- | --- |
|  | PZ-CG | CG-MUS | PZ-MUS | PZ-CG | CG-MUS | PZ-MUS |
| **Abstinence** | 1.60 (0.31) | 3.77 (0.76) | 5.88 (0.56) | 1.63 (0.35) | 3.65 (0.66) | 5.82 (0.95) |
| **Ejaculation** | 1.26 (0.20) | 3.04 (0.54) | 3.76 (0.56) | 1.24 (0.21) | 2.94 (0.50) | 3.57 (0.51) |
| **$P$-value** | 0.000 | 0.000 | 0.000 | 0.000 | 0.000 | 0.000 |

*Data are presented as mean (SD).*

**Fig 1.**
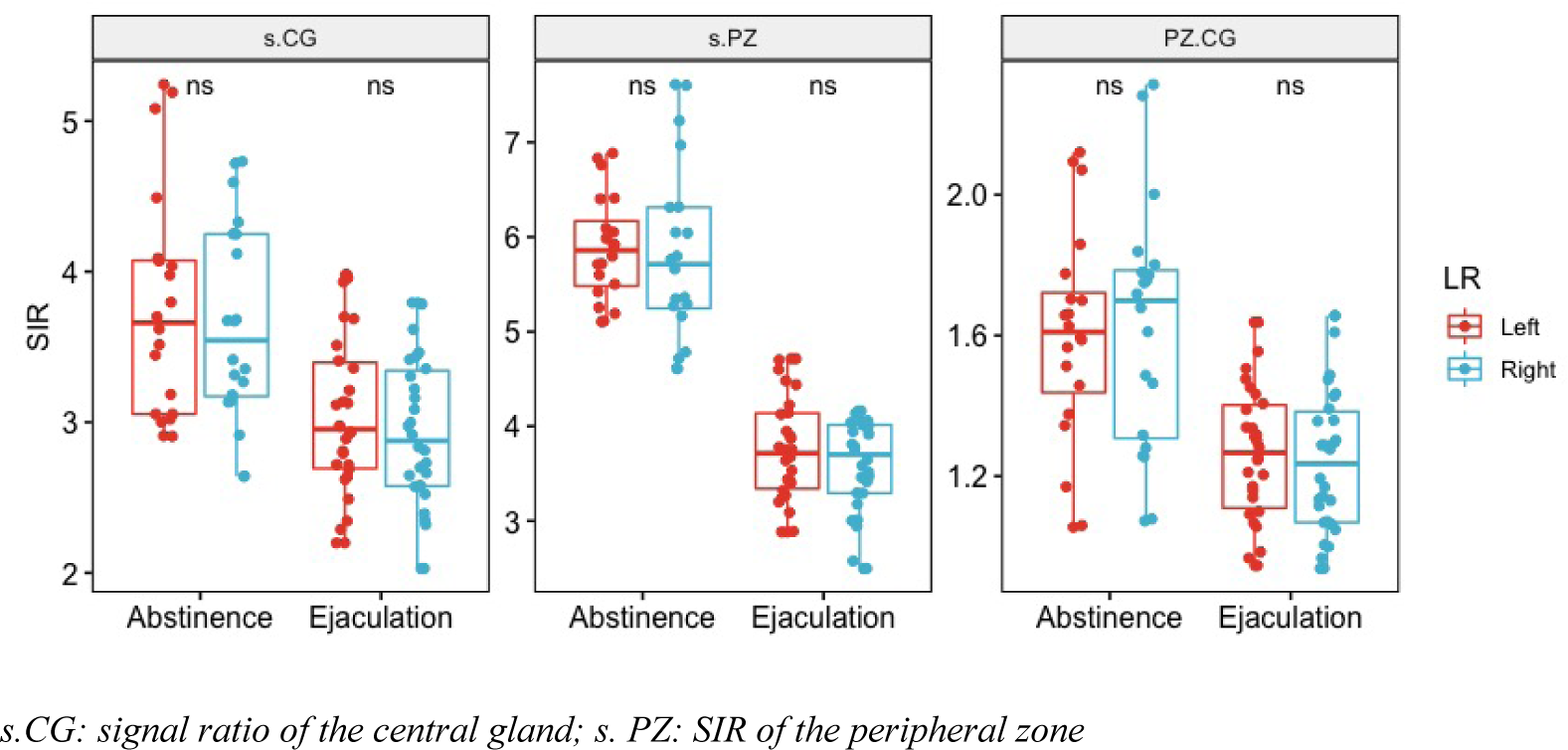
The SIR of the CG, PZ, and the PZ-CG between left and right sides. There are no difference between the left and right sides in the same group, *P*>0.05.

**Fig 2.**
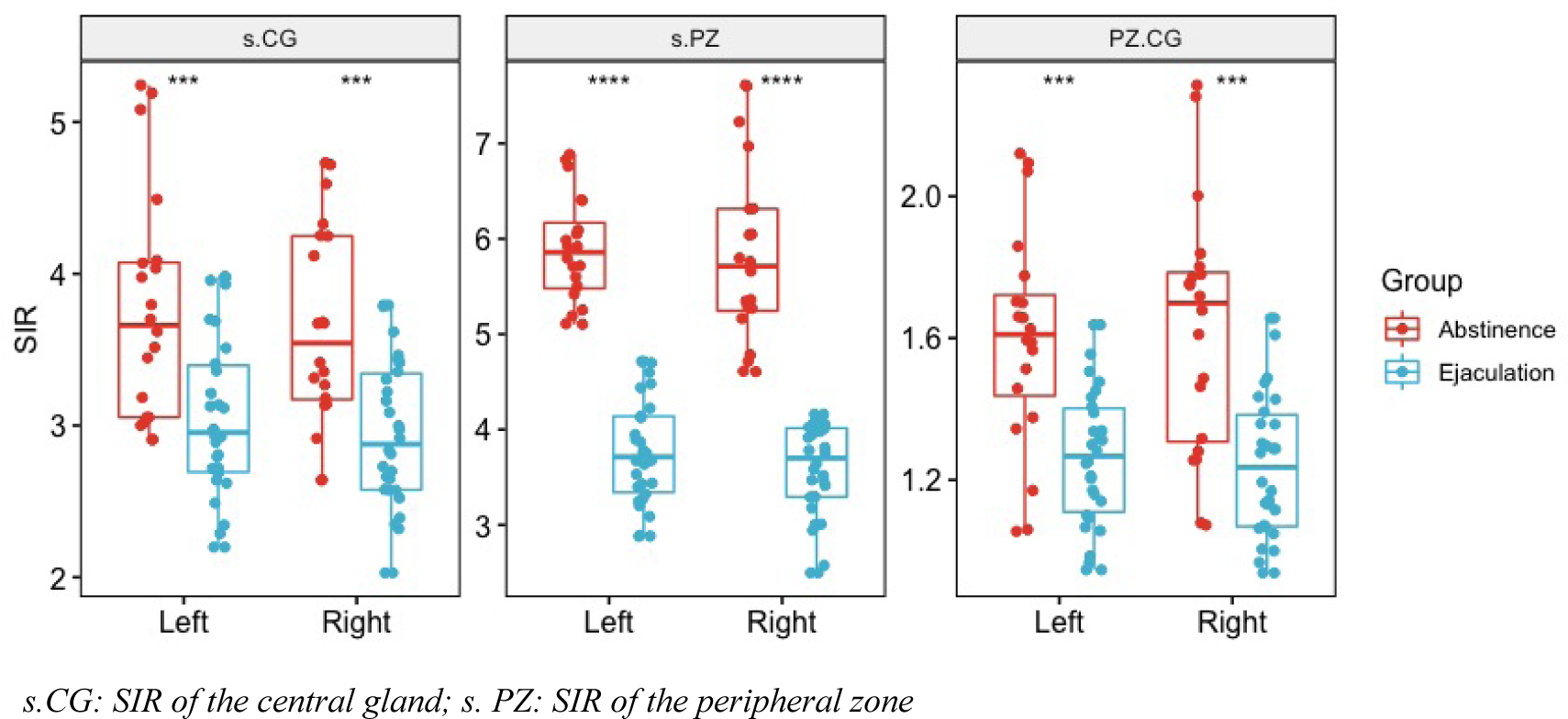
The signal ratio of CG, PZ and PZ-CG on the same side between the two groups. The SIR of CG, PZ and PZ-CG on the same side between the abstinence group and ejaculation group were statistically different, all *P*=0.000.

## DISCUSSION

By comparing the T2WI of two groups, our study shown that the structure of the prostate gland in the abstinent group was easier to identify through qualitative analysis. Moreover, quantitative analysis shown that the signal of peripheral zone and central gland were increased in abstinence group, and the contrast between peripheral zone and central zone was more obvious.

Previous studies have shown that ejaculation may affect the T2WI and ADC signals of the prostate. Medved et al. and Shin et al. manually drawn ROIs at a typical layer in their study, and segmented the CG and PZ of the prostate completely which is mainly based on their clinical experience ^[11-12]^. But the young males have a relatively small volume of TZ, it is hard in zonal differentiation, especially the signal of prostate would be changed after ejaculation. These would be a difficult challenge for manual segmentation. Due to this reason, Barrett *et al*. didn’t segment the different zone, they just outlined entire prostate as a ROI, but the different structures of the prostate did have different features in T2WI, this may affect the results^[13]^. In order to avoid these impacts, we were not looking for a completely separation of the prostate into CG and PZ. We just outlined the same size ROIs on both sides of the prostate gland,they were located in the CG and PZ on the left and right sides respectively. Our quantitative analysis about the left and right SIR in same group shown that there is no statistical difference between them. It reflects that the ROI in the bilateral contralateral position belongs to the same structure of the prostate. It demonstrated that our method of outlined ROIs is more reasonable.

In our study, we found that the SIR of PZ and CG were lower in the ejaculation group than those in the abstinence group, and the reduction of SIR of PZ was more obvious. This indicated that the PZ and CG were more easily distinguished in the abstinence group. About 70% of prostate glands are located in the PZ of the prostate, and 25% are located in the CZ. The glands slowly produce prostatic fluid and store it in the glands, therefore, the prostate present as a hyperintense. The liquid is an important component of semen, accounting for approximately 30%. Prostatic fluid from different areas is discharged into the urethra during ejaculation. We observed that in this study, the prostate signal is decreased after ejaculation. As the peripheral zone stored more prostatic fluid, the signal of the peripheral zone decreased more than that of the central zone after ejaculation.

As we known, prostate cancer appears as a hypointense lesion on T2WI due to the destruction of glandular structure ^[14]^. We hypothesized that decreased prostatic signal after ejaculation may affect the detection of prostate malignancy, especially using some quantitative methods. Therefore, a period of abstinence before MRI examination is necessary. However, PI-RADs specifically points out that abstinence for 3 days comes from clinical experience and there is no significant evidence to support this view. Through this retrospective study, patients were divided into two groups according to whether they abstained for 3 days or not, the above results were observed, which support the recommendation of abstinence before examination. However, it is necessary to further investigate the reasonable duration of abstinence by comparing MRI images at multiple time points.

There are still some limitations in this study. First, the study was conducted on healthy young men. However, the clinical situation is more complicated. Especially, with the increase of age, the prostate volume and the T2W signal intensity of the PZ was also increases. How does it changes in the enlarged prostate after ejaculation has not been studied. In particular, how does the signal intensity change after ejaculation in prostate cancer patients need further research and analysis. Moreover, since it is difficult and inaccurate to completely outline ROIS manually, this study adopts the method of selecting points on both sides of the region to outline ROIS, which may lead to bias of the results. With the development of AI technology, it is possible to use machine learning method to completely automate the division of the prostate, which may avoid the bias caused by manual division. Moreover, PI-RADS recommends mp-MRI containing T2WI, DWI, DCE and other sequences for the detection of prostate cancer. In this study, due to the limitation of research data, we just analyzed the difference of T2WI. Therefore, it is necessary to conduct further study on other sequences to analyze the possible influence of ejaculation on them.

## CONCLUSION

In conclusion, we find that the T2WI-SIR of prostate, both the CG and PZ, were decreased after ejaculation in healthy young male. And the SIR of PZ decreased more than CG, this makes it more difficult to identify the internal structure of the prostate. From these results, we provide some evidence of abstinence prior to the prostate MR examine. As PI-RADS recommend,we support that the abstinence time before prostate MRI is at least 3-days.

## Data Availability

All relevant data are within the manuscript and its Supporting Information files.

## REFERENCES

[1] Jemal A, Bray F, Center MM, et al. Global cancer statistics. CA Cancer J Clin. 2011;61: 69–90.

[2] Sung H, Ferlay J, Siegel RL, et al. Global cancer statistics2020: GLOBOCAN estimates of incidence and mortality worldwide for 36 cancers in 185 countries[J]. CA Cancer J Clin, 2021, 71(3):209–249.

[3] Zeng H, Zheng R, Guo Y, et al. Cancer survival in China, 2003-2005: a population-based study[J]. Int J Cancer, 2015, 136(8):1921–1930.

[4] Zeng H, Chen W, Zheng R. Changing cancer survival in China during 2003-15: a pooled analysis of 17 population-based cancer registries[J]. Lancet Glob Health, 2018, 6(5):555–567.

[5] Rouviere O, Moldovan PC. The current role of prostate multiparametric magnetic resonance imaging[J]. Asian J Urol, 2019, 6(2):137–145.

[6] Richenberg J, Løgager V, Panebianco V, ert al. The primacy of multiparametric MRI in men with suspected prostate cancer[J]. Eur Radiol, 2019, 29(12):6940 –6952.

[7] Habchi H, Bratan F, Paye A, et al. Value of prostate multiparametric magnetic resonance imaging for predicting biopsy results in first or repeat biopsy. Clin Radiol 2014;69(3): e120–e128

[8] Baris Turkbey, Andrew B Rosenkrantz, Masoom A Haider, et al. Prostate Imaging Reporting and Data System Version 2.1: 2019 Update of Prostate Imaging Reporting and Data System Version 2[J]. Eur Urol. 2019, 76(3):340–351.

[9] Milica Medved, Steffen Sammet, Ambereen Yousuf, et al. MR imaging of the prostate and adjacent anatomic structures before, during, and after ejaculation: qualitative and quantitative evaluation[J]. Radiology, 2014, 271(2):452–460.

[10] Jeffrey C Weinreb, Jelle O Barentsz, Peter L Choyke, et al. PI-RADS Prostate Imaging - Reporting and Data System: 2015, Version 2[J]. Eur Urol, 2016, 69(1):16–40.

[11] Takeshi Shin, Yasushi Kaji, Toshiro Shukuya, et al. Significant changes of T2 value in the peripheral zone and seminal vesicles after ejaculation[J]. Eur Radiol, 2018, 28(3):1009–1015.

[12] Milica Medved, Fatma N Soylu-Boy, Ibrahim Karademir, et al. High-resolution diffusion-weighted imaging of the prostate[J]. AJR Am J Roentgenol, 2014, 203(1):85–90.

[13] Tristan Barrett, Soleen Ghafoor, Rajan T Gupta, et al. Prostate MRI Qualification: AJR Expert Panel Narrative Review[J]. AJR Am J Roentgenol, 2022, 219(5):691–702.

[14] Tiago Oliveira, Luís Amaral Ferreira, Carlos Miguel Marto, et al. The Role of Multiparametric MRI in the Local Staging of Prostate Cancer[J]. Front Biosci (Elite Ed), 2023, 15(3):21.

